# Local wastewater monitoring as a complementary tool in a mumps outbreak investigation in the Netherlands: a proof-of-concept study

**DOI:** 10.1101/2025.08.05.25333031

**Authors:** Maja Joosten, Aart C. Dijkstra, Maarten de Jong, Remy P.S. Schilperoort, Jeroen Langeveld, Marlous Prins, Gregorius J. Sips, Ewout Fanoy, Miranda de Graaf, Gertjan Medema, Loes Jaspers

## Abstract

**Background:** A recent outbreak of mumps resulted in a concerningly low number of classical disease notifications, causing a search for alternative methods of monitoring.

**Aim:** This study aims to evaluate whether mumps virus RNA can be detected in wastewater, and how these measurements relate to clinical monitoring.

**Methods:** Suspected mumps cases were recorded by local GP practices (n = 13) for 20 weeks and laboratory-reported notifications were collected. Social media monitoring was conducted retrospectively. Local wastewater monitoring using passive samplers was deployed downstream of the house and school of a confirmed case, in wastewater pumping stations serving the confirmed case and neighbouring towns, in the downstream pumping? station serving the entire area, and in a pumping station outside the affected area as a control. Wastewater samples were analysed using RT-PCR.

**Results:** GPs reported 50 suspected patients, of which 24 were confirmed. Several GPs noted suspected underreporting in the area.

The passive sampler near the confirmed case detected virus RNA directly post-diagnosis, matching the genotype from patient material, followed by a negative signal afterwards, while the sampler at the school and the local pumping station detected rising concentrations as more cases were reported. Samplers in neighbouring towns suggested an active outbreak, including positive measurements in a nearby town with no reported cases. Control samples were negative.

**Conclusion:** This study demonstrates the first successful proof-of-concept for local wastewater monitoring of mumps and indicates a correlation between wastewater and clinical monitoring. These results suggest a role for local wastewater monitoring using passive samplers in creating situational awareness during an outbreak, even when cases are not reported.

## Introduction

Mumps (parotitis epidemica) is an acute infectious disease caused by mumps virus, a member of the family *Paramyxoviridae* (1). It is a moderately to highly contagious infection that is restricted to humans. Transmission occurs by inhalation of respiratory droplets from infected individuals in their infectious period (2). The incubation period ranges from 15 to 24 days (median 19 days) and infected patients are contagious from two days prior to the onset of clinical symptoms to five days afterwards (3). Mumps is characterised by swelling of the parotid gland, either unilateral or bilateral, which occurs in 60 - 70% of all infections and 95% of patients with symptoms (3). While the disease is usually mild, it can be accompanied by complications, such as aseptic meningitis, deafness, encephalitis, orchitis and oophoritis (3). The former three complications are primarily seen in unvaccinated children, while the latter two are seen post-pubescent (3).

Mumps has previously been the cause of significant morbidity worldwide; 40 - 726 cases per 100.000 individuals per year (4). The incidence of mumps has decreased significantly since the introduction of the vaccine, showing a 100-fold decrease between pre- and post-vaccine eras in several European and Asian countries(4). The combination vaccine against measles, mumps and rubella (MMR) was first introduced in the Netherlands in 1987 for all children aged 14 months and 9 years as part of the Dutch National Immunisation Program (5). The disease had since been virtually eliminated for several years, until an epidemic among unvaccinated children in 2007 (1,5) was cause for renewed surveillance, followed by an epidemic among vaccinated students between 2009 and 2013 (5,6). Since then, between 39 and 131 cases have been reported annually, with a sharp decline during the COVID-19 pandemic (7).

In November 2023, an outbreak of mumps occurred in a region of the Netherlands with low vaccination coverage (8). It was suspected that the outbreak was much larger than the formally notified number of cases suggested, as diagnostic confirmation may be performed infrequently, and suspected cases are not notifiable. Additionally, general practitioners in the area reported informal signals of possible cases that did not come in for consultation. Therefore, insight into the true scope of the outbreak was lacking, which complicates the weighing of interventions. This prompted the search for alternative methods of monitoring besides the established notification system of the Dutch Public Health Act (9–11). In other outbreaks, social media monitoring has been applied. Furthermore, monitoring of infectious diseases in wastewater has been extensively developed and implemented during the COVID-19 pandemic (12), and multiple agents have been studied and can be observed in wastewater (13). To the authors’ knowledge, no studies have assessed whether mumps virus can be detected in wastewater and how this relates to the prevalence of mumps.

Mumps can be detected in urine in the first week after disease onset using PCR (3). This suggests that wastewater monitoring could be an efficient tool as an early warning system and for epidemiological monitoring of mumps (12). Wastewater monitoring has the added advantage of covering not only symptomatic individuals who have been tested, but also those who have not been tested, thereby limiting the problem of underreporting or underdiagnosis (12,14). However, wastewater monitoring might also detect virus shedded by asymptomatic cases which would not be notifiable and, whilst providing an insight into the presence of infections, may not directly translate to the presence of notifiable cases within a community.

Typically, wastewater monitoring has been conducted at large wastewater treatment plants, which are equipped with (costly) flow proportional 24-hour sampling. Installing such devices is not easily applicable to small catchment areas. Passive samplers present a low-cost, safe and easy alternative to traditional wastewater sampling within the wastewater catchment. They offer an advantage for small scale sampling with higher spatial resolution, making local and on-demand monitoring possible (15,16).

Passive sampling involves the deployment of a device in a sewer system for 24 up to 72 hours, allowing for infectious agents in wastewater to interact with the absorbent in the passive sampler. At the end of the deployment, the passive sampler is analysed for the presence of specific pathogens. Deployment of passive samplers in water systems is relatively straightforward (i.e., no specialized skills required), rapid, and usually does not require confined space entry permits. Furthermore, the continuous exposure of the passive sampler to wastewater reduces the sampling errors that exist when taking discrete samples.

The aim of this study was to identify whether mumps virus could be detected in wastewater samples during an outbreak of mumps in the Netherlands, using PCR testing on passive sampler material. Furthermore, we aimed to evaluate the association between wastewater findings of mumps and clinical monitoring of mumps cases to gain insight in the usefulness of the method for outbreak response by improving situational awareness.

## Methods

### Setting and design

In this proof-of-concept study, we prospectively collected confirmed, probable, and possible cases of mumps between November 1^st^ 2023 and March 31^st^ 2024 from established and impromptu clinical surveillance systems, and related this data to the PCR results of wastewater monitoring using passive samplers. The study was conducted in the North-East subregion of Gelderland, a province in the Netherlands. This subregion consists of several villages with tightly connected communities. The vaccination coverage indicated great interregional diversity. For example, while one municipality has a global vaccination rate of 74% (17), the local rate varies between 33% and 88% for specific subregions (unpublished data).

### Clinical surveillance systems

In the Netherlands, the Dutch Public Health Act requires doctors, laboratories, and (health care) institutions in the Netherlands to notify the Municipal Health Service (MHS) in their region when designated infectious diseases are diagnosed or suspected (9). Mumps is one of these notifiable diseases (10). This system typically requires patients to be seen by a physician and undergo diagnostic testing before cases are notifiable. Notifications for mumps in the area of one municipal health service (MHS Northern- and Eastern-Gelderland) were collected for the duration of the outbreak. Due to suspected underreporting, manual syndrome surveillance was added to the clinical data collection procedure. The number of possible and confirmed cases per GP practice was gathered using weekly phone calls and an Excel sheet for all local GP practices in the region (n = 13).

### Case definitions

A confirmed case was defined as an individual with at least one of the following three symptoms: (i) acute onset and painful swelling of the parotid or other salivary gland, (ii) orchitis and (iii) meningitis; as well as meeting at least one of the two following criteria: (i) laboratory-confirmed infection with mumps virus or (ii) contact within 4 weeks prior to disease onset with a confirmed case; following the definition of a notifiable case according to the Dutch Public Health Act (9,18). Probable cases were defined as individuals with the same symptoms but lacking a laboratory confirmation or epidemiological link.

Possible cases were defined as any individual fitting the clinical presentation of mumps, who consulted their GP for a mumps-related consultation and of whom the GP made a registration in their electronic patient file using the ICPC-code D71 ‘mumps’ (19). The latter were reported as aggregated counts per GP practice, while the former two categories were gathered as individual cases from notification data.

### Social media monitoring

Social media monitoring was conducted retrospectively using Coosto (20). This social media monitoring-tool enables systematic searches of online content in different online sources. The tool keeps an archive of all Dutch posts published on social media and thereby provides the opportunity to extract all posts on a certain topic, providing the time and date of the online messages. A search was conducted using the keywords ‘mumps’ or ‘swelling of cheeks’ in combination with ‘MMR’ or ‘vaccine’ or ‘infection’ or ‘outbreak’ during the period of November 1^st^ and March 31^st^.

### Wastewater monitoring

Passive samplers were deployed at several regional points of interest. To confirm that mumps virus RNA can be detected in wastewater, samplers were installed in the sewers downstream from a confirmed case after disease onset. Similarly, passive samplers were placed near the involved primary school, as well as in pumping stations in sewer districts with confirmed or suspected cases (towns A and B, see Figure 1). Pumping stations in neighbouring districts (towns C and D) were sampled to monitor the spread of the epidemic. Town E was monitored at the beginning of the main gravity transport sewer to the downstream wastewater treatment plant, as there is no upstream junction or pumping station available that encompasses the entire town without the addition of wastewater from the other (upstream) districts. Passive samplers were deployed five weeks after the first case was reported and sampling was repeated until March 28th at one- to four-week intervals. Control samples were obtained from a region in Rotterdam, The Netherlands, where no notified mumps cases were present at the time of sample collection. Sewage flow rates were determined from standard process logs in the pumping stations, with a monitoring frequency of 2 minutes.

**Figure 1.**
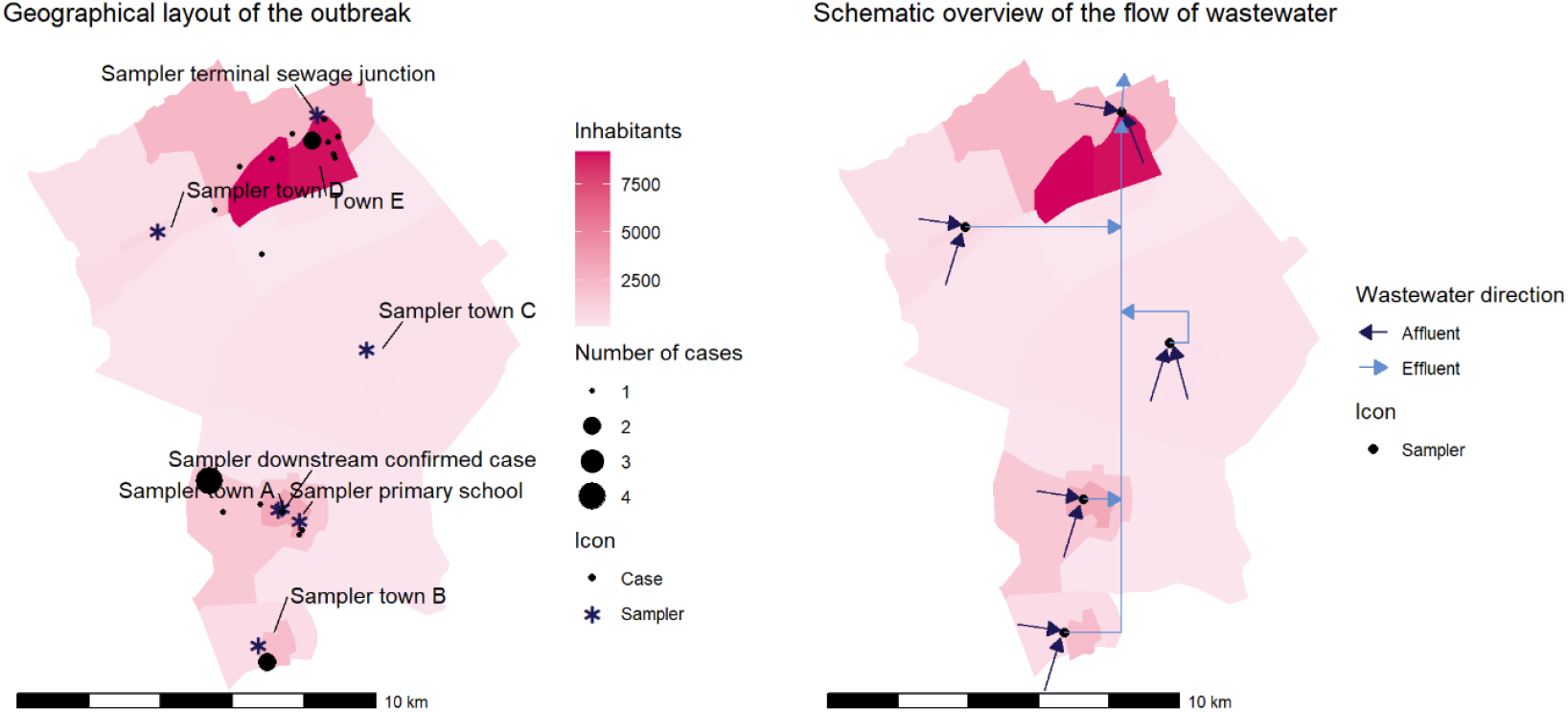
Geographical layout of samplers and cases (left) and a schematic overview of the sewage network (right). The involved towns are located in a rural area. They are geographically distinct and separated by uninhabited regions. Wastewater flows in the northeast direction, combining roughly at the end of each town, where passive samplers were placed. The network converges underneath town E and empties at the terminal sewage junction. Therefore, town E could not be separately sampled.

Passive samplers consist of a 3D printed porous plastic container filled with absorbent materials, e.g. cotton tips and membranes (14,15). Passive samplers were placed manually at the points of interest for an exposure period of 48 hours, after which the materials were collected and transported to the laboratory.

### PCR testing and sequencing

Membranes were eluted and then processed in nucleic acid extraction buffer (Nuclisens, Biomerieux, Amersfoort, the Netherlands), cotton-tips were directly placed in nucleic acid extraction buffer (Nuclisens, Biomerieux, Amersfoort, the Netherlands), and processed as described previously (21). RT-PCR testing was performed on the sample extracts targeting the F-gene as described previously (22). Reactions were considered positive if the cycle threshold (Ct) was below 40 cycles. Readings above 40 cycles were discarded.

PCR crAssphage and human *Bacteroides* (HF183) were used as an indicator of the amount of human fecal matter in the wastewater and thus its dilution (23). These are used to determine the dilution rates during storm events and serve as an indicator for the quality of the sampling. Low numbers could indicate blockage of the passive sampler, for example by sanitary towels, and thus a less representative sample.

Typing of the mumps virus was performed by sequencing the small hydrophobic (SH) gene from the same sample extract as previously described (24), followed by BLAST analysis of the nucleotide sequence to determine the genotype.

### Data analysis

Comparative analyses were performed to determine the trend between wastewater measurements and clinical findings. Wastewater measurements were plotted against confirmed, probable and possible cases to assess this interaction. Negative readings were visualized using a Ct-value of 42. Direct quantification of the Ct-values compared to disease incidence was not possible, due to a lack of reliable incidence metrics. Finally, sequencing results of clinical cases and wastewater samples were compared in order to establish a connection between the underground and aboveground findings. All analyses were performed in R (version 4.3.3, The R Foundation).

### Ethical considerations

This study was conducted in the context of outbreak control, as outlined in the Dutch Public Health Act. The study did not require approval from the medical ethical committee, as it does not concern medical scientific research as defined by the Dutch Central Committee on Research Involving Human Subjects. This was independently confirmed by the Medical Research Ethics Committee (METC NedMec). Wastewater monitoring was applied with approval of the involved municipalities and at catchment areas covering at least 50 addresses, ensuring that individual persons could not be identified.

## Results

### Description of cases

A total of 24 confirmed and probable cases with symptom onset between November 1^st^, 2023, and March 31^st^, 2024, were registered (Figure 2), with the first confirmed case being reported on the 16^th^ of November. Three individuals (12.5%) developed complications, all had orchitis. A total of 15 cases showed an epidemiological link with other mumps cases (Table 1).

**Table 1.**
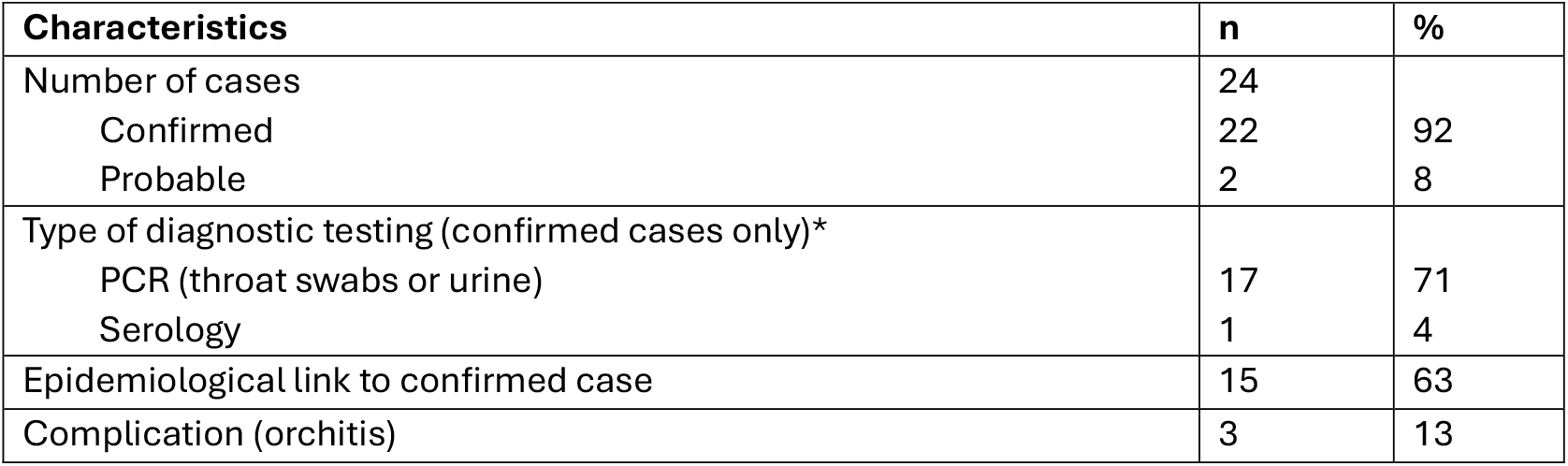
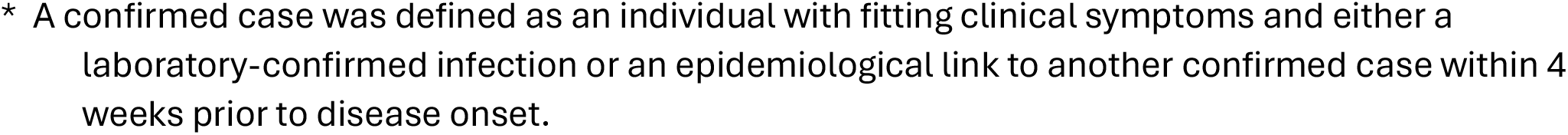
Overview of the probable and confirmed notified cases.

| Characteristics | n | % |
| --- | --- | --- |
| Number of cases | 24 |  |
| Confirmed | 22 | 92 |
| Probable | 2 | 8 |
| Type of diagnostic testing (confirmed cases only)* |  |  |
| PCR (throat swabs or urine) | 17 | 71 |
| Serology | 1 | 4 |
| Epidemiological link to confirmed case | 15 | 63 |
| Complication (orchitis) | 3 | 13 |

\* A confirmed case was defined as an individual with fitting clinical symptoms and either a laboratory-confirmed infection or an epidemiological link to another confirmed case within 4 weeks prior to disease onset.

**Figure 2.**
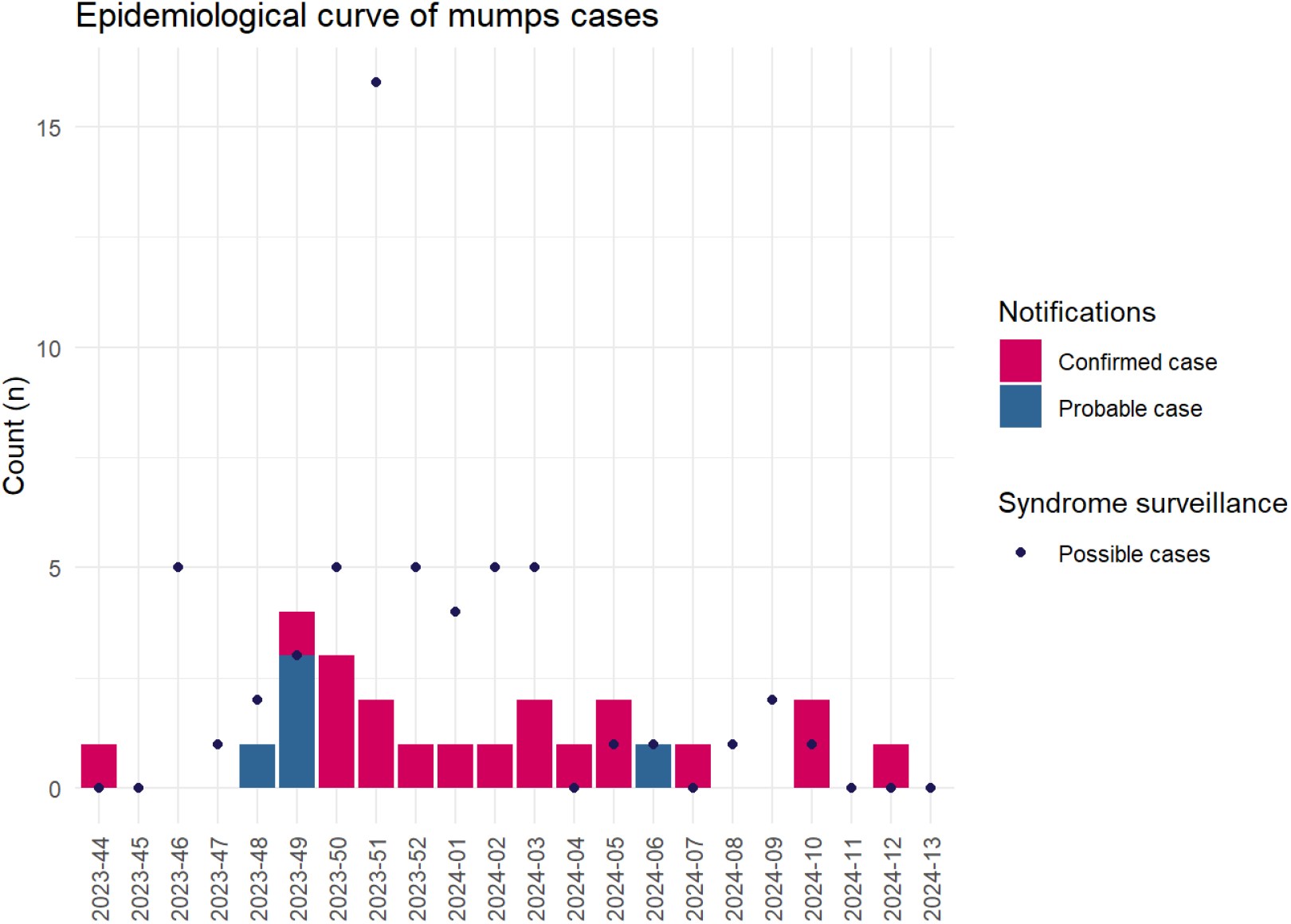
Number of probable and confirmed cases and GP reported possible cases in the affected region in the northeast of Gelderland between week 44 2023 and week 13 2024.

GPs reported a total of 57 possible cases fitting the clinical description of mumps. The highest number of new possible cases was found in week 51 (Figure 2). These cases were found in the town of a confirmed case (Town A, see Figure 1) in the first five weeks of monitoring, and subsequently in two of the surrounding towns (Town B+E). No possible cases were reported, and no notifications were received from town C. Several GPs noted likely underreporting in the area, based on informal signals, indicating that the true number of cases was likely higher. In the last three weeks of monitoring no possible, probable or confirmed cases of mumps were reported.

### Social media monitoring

A total of 96 messages were found on social media in the time period November – March. The majority were messages on X (52 messages), followed by 20 messages from news sources, twelve from blogs, ten from Facebook and two from Youtube. All messages were on the topic of MMR vaccination in general; there were no messages addressing the mumps epidemic. The number of messages did not increase during weeks with more mumps cases.

### Wastewater monitoring results and clinical findings

Mumps virus RNA was found in wastewater samples using passive samplers. Wastewater monitoring indicated patterns consistent with clinical surveillance data. The first sampler deployed downstream of a confirmed case (Figure 3) detected mumps virus RNA particles 12 days after disease onset (Ct-values 35.9 and 37.9 for cotton tips and membrane, respectively), followed by a sharp decline and subsequent negative signal for several weeks afterwards. The community samplers at the school and the local pumping station (Town A) initially yielded a rising concentration of mumps virus RNA coinciding with an increase in reported cases, and tested negative for several weeks afterwards, coinciding with a lack of reported cases. Samplers at pumping stations in neighbouring towns (Town B and D) showed varying levels of mumps virus RNA over time, roughly matching clinical surveillance data (Figure 3). The sampler in Town C yielded positive measurements while no possible cases were reported by GPs and no notifications were received in that area.

**Figure 3.**
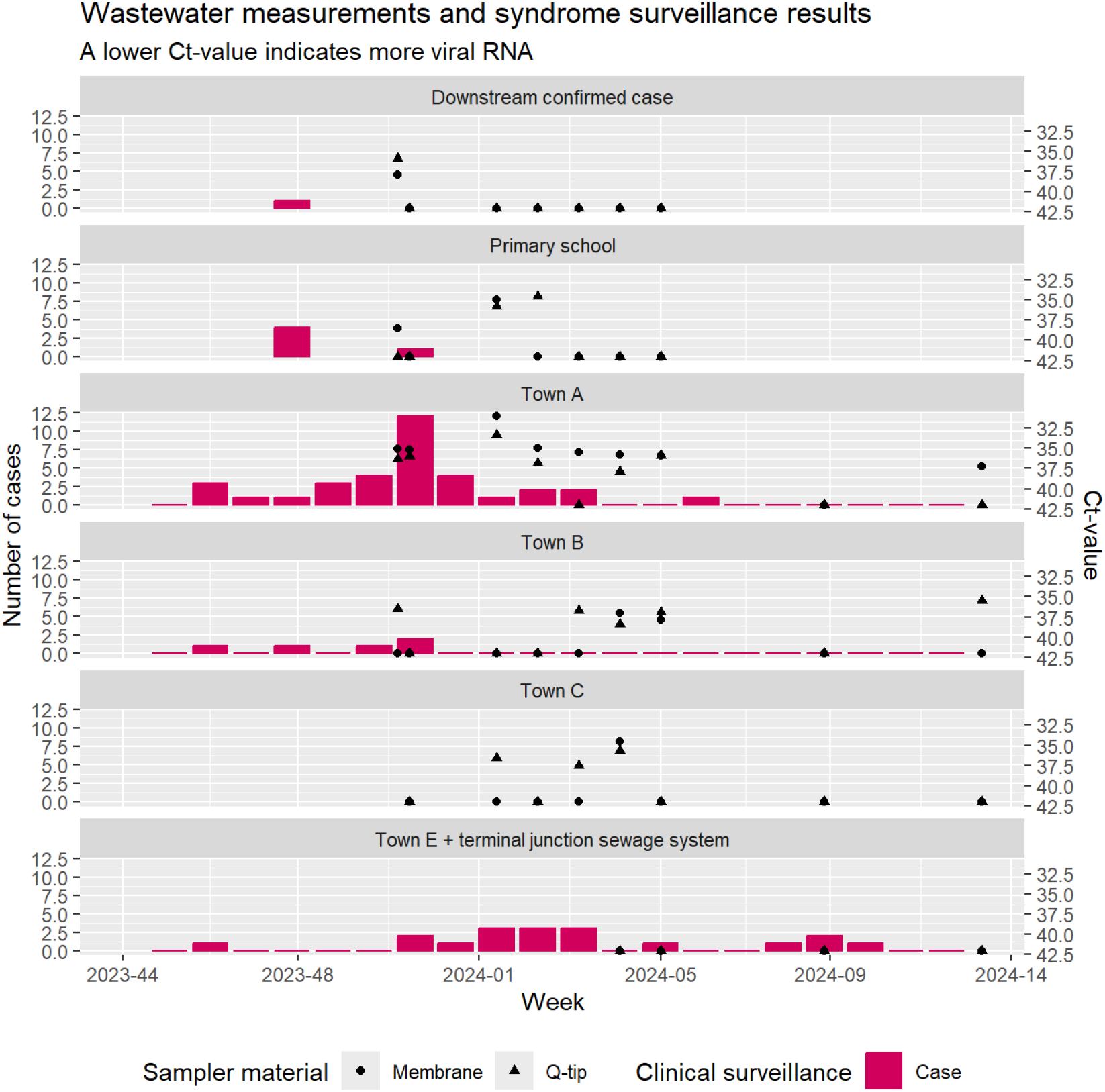
Observed trend between wastewater measurements and possible, probable or confirmed cases per catchment area. Town D and control measurements not shown; all measurements were negative. N.B.: Negative results are plotted as having a Ct of 42 for visibility, while in reality values above 40 were not reported.

Notably, the sampler deployed at the station furthest downstream showed negative signals, while those upstream closer to cases showed positive signals. Control samplers from a region without known mumps cases were negative.

#### Wastewater flow rates in relation to DNA particle concentration

In a week with heavy rainfall a gross total of 12,000m^3^ sewage and rainwater was pumped through the pumping station in town A in 48 hours, while in a week without heavy rainfall a total of 4,600 m^3^ sewage and rainwater pumped through the sewage station in 48 hours. This dilution of a factor 2-3 by rainwater had little effect on the measured concentrations of crAssphage and HF183 (Figure 4). If the rainfall diluted the mumps virus signal, the observed Ct-value of week 1 in Town A (Figure 3) could have maximally been 1 Ct lower.

**Figure 4.**
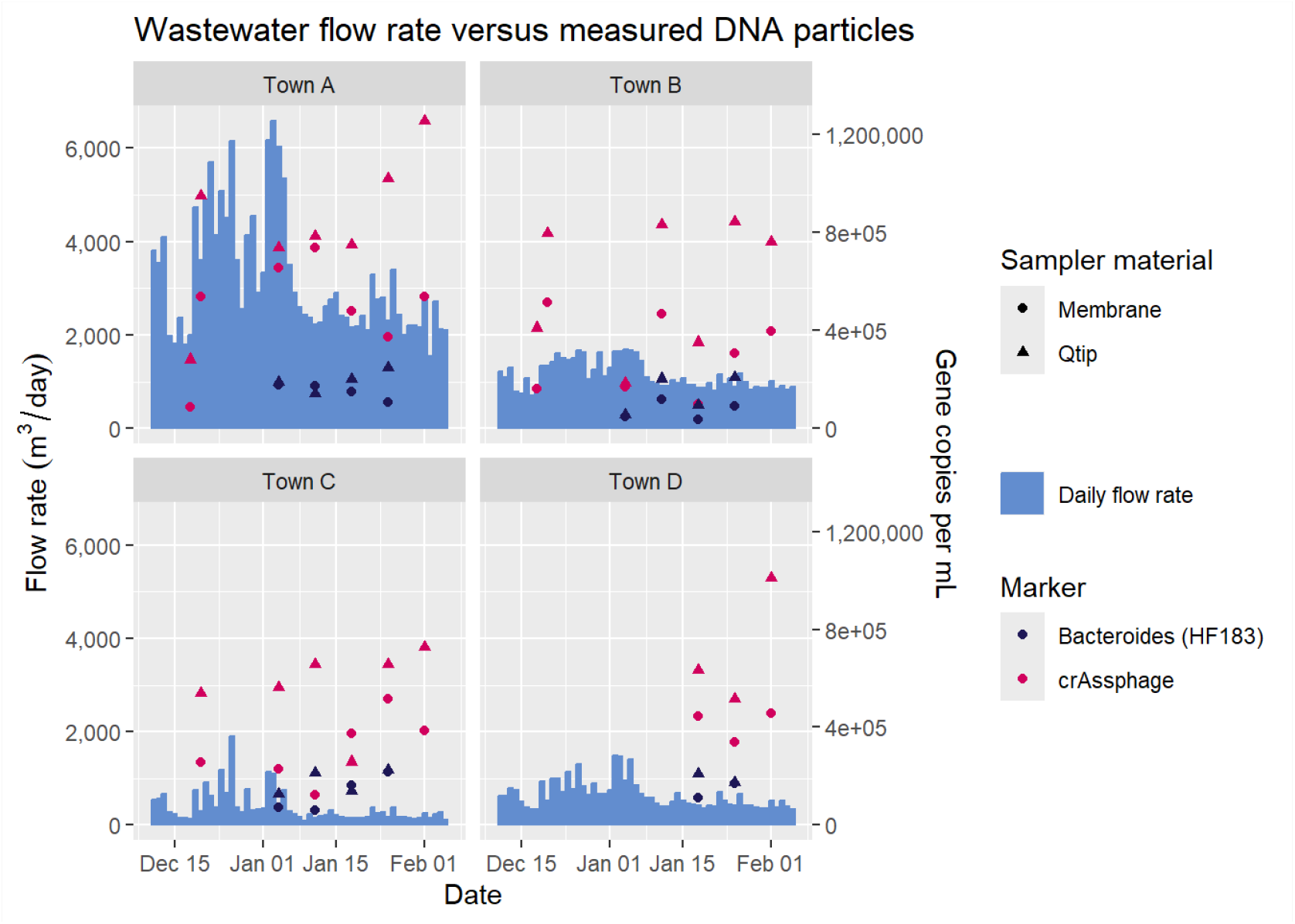
Wastewater flow rate and measured DNA particles for human *Bacteroides* (HF183) and crAssphage during a period of heavy rainfall (left hand side of all graphs) and light to no rainfall (right hand side).

### Sequencing

The SH gene was successfully sequenced from samples collected from the sampler downstream of the index patient, the sampler downstream from the school, and the samplers at locations A & C. Mumps genotype G was identified in both wastewater and clinical isolates. The SH gene sequences obtained from wastewater and the patient were identical. Analysis showed that this sequence differed from those associated with other mumps outbreaks and clusters reported in the Netherlands in 2023 (sequence MuVs/Gelderland.NLD/45.23, available in Genbank).

### Use of sewage data for public health response

Based on the additional information from wastewater monitoring, particularly regarding the presence of the virus in surrounding villages (Town B and C) over an extended period, the MHS decided to inform the local government of the ongoing mumps outbreak. Subsequently, using this information, the government informed the citizens about the circulation of mumps, its symptoms, and the measures they could take to protect themselves.. Furthermore, the MHS implemented measures to enhance syndrome surveillance at general practitioners’ offices for future public health responses. Although the MHS considered a vaccination campaign based on wastewater monitoring, it ultimately decided against it due to the religious context of this specific area. Had the outbreak occurred in a different context, a vaccination campaign might have been a viable option based on these wastewater monitoring results.

## Discussion

This proof-of-concept study indicates that mumps virus RNA can be detected in wastewater using passive samplers when mumps is present in the community. To the author’s knowledge, this is the first study applying this technique – which is well established for other viruses such as the SARS-CoV-2 virus – in a community setting with an outbreak of mumps. Mumps virus RNA was found in several locations with confirmed cases, while the control location showed no measurable virus RNA concentrations. Furthermore, identical genotypes of mumps virus RNA were found in wastewater and patient samples. This genotype is different than other recent outbreaks of mumps in other regions of the Netherlands.

### Trends and silent transmission

A trend was observed between the results of wastewater testing and reported cases. The first sampler, deployed downstream of a confirmed case, yielded positive results 12 days after disease onset, and negative results 29 days after disease onset. This suggests that a mumps infection yields detectable virus RNA concentrations in wastewater for a short period after disease onset. The wastewater samples were positive in close proximity of the school with a suspected outbreak, and in town A (town with the suspected outbreak) as the reported number of cases increased. Consecutively, the samples near the confirmed case and the school came back negative when the number of cases subsided. Interestingly, the samples in town A remained positive in the weeks thereafter, even whilst reported cases decreased, possibly implying ongoing silent transmission in the town. A similar pattern was observed for town B. In town C, wastewater samples came back positive slightly later in time, whereas no cases were reported. This could imply some unreported geographical spread of the outbreak to towns in close proximity of the outbreak.

Samples from the control location were negative, indicating that mumps virus RNA is not usually found in wastewater. This is of importance, as it was unknown whether recently vaccinated individuals emit mumps virus RNA in wastewater, as MMR is an attenuated viral vaccine. In the specific region of this study, the last vaccination campaign for the MMR vaccination was a few months prior to the start of the wastewater monitoring. Both the timeframe and the negative control samples make it highly unlikely that MMR vaccination could be a source of the positive wastewater samples that were found in this study.

### Underreporting

Mumps infections may be prone to underreporting, as symptoms are often mild and diagnostic testing may be performed infrequently (25,26). First, individuals with mumps may not contact their GP, or only when they have complications. In this study, the number of reported cases (n = 24) was significantly lower than the number of possible cases (n = 57) based on manual syndrome surveillance, suggesting that many patients did not come in for consultation. As for complications; this study found a complication rate of 13%, which is in line with literature (18). Second, GPs often diagnose mumps clinically without any further laboratory testing. Third, around 33% to 63% of people with mumps remain asymptomatic, depending on vaccination status (18) The benefit of wastewater monitoring is that it could be used as an indicator of transmission of mumps regardless of diagnosis- or treatment-seeking behavior. Local wastewater monitoring could thus be of added value for situational awareness during an outbreak with suspected underreporting, allowing the MHS to inform municipalities, GPs, and residents about potential measures in due time. During this outbreak, the additional information from wastewater monitoring was used by the municipality to inform the public about the outbreak.

### Local situational awareness

In town E, the virus was not picked up by the passive samplers, even though six cases were reported in this town. However, town E could not be isolated using the sewer system. Samplers were thus deployed further away from the location of the cases at a beginning of the main transport sewer, meaning shedded virus particles could have been more diluted with domestic wastewater and rainwater. In contrast, the passive sampler in town C showed several positive signals, despite a lack of reported cases. This increased geographical spread would thus be unknown if passive samplers had not been deployed. Consequently, wastewater monitoring with passive samplers in close proximity to a suspected outbreak could help create local situational awareness or could be used complementary to more established periodic (national) wastewater surveillance, which are implemented for larger catchment areas (27). Local wastewater monitoring with passive samplers was recently proven to be effective for monitoring other agents, such as polio- and measles virus (28), hepatitis A virus (21), and SARS-CoV-2 virus. This study indicates that it can be used for the detection and monitoring of mumps virus as well.

### Methodological considerations

Several limitations need to be taken into consideration. First, it was unknown how many cases were present in the measurement period, due to suspected underreporting. Attempts to address this issue by including ‘possible cases’ from the automated GP syndromic surveillance of the Netherlands Institute for Health Services Research (NIVEL) were unsuccessful, as there was no representative surveillance station within the subregion of interest for this study. Therefore, manual syndromic surveillance was implemented by making weekly phone calls to the involved GP practices.

Second, since the true number of cases is unknown, quantification of these results is impossible in this study.

## Conclusion

This study demonstrates the first proof-of-concept for local wastewater monitoring of mumps virus and indicates a trend between wastewater and clinical monitoring. The results suggest a complementary role for passive samplers in identifying and monitoring the course of an outbreak and creating situational awareness, especially when cases are not or underreported via established surveillance systems. Such information can aid public health decision making and direct the implementation of localized preventative interventions such as vaccination strategies, public health communication and advice to local governments.

Future studies could focus on further quantifying the association between wastewater testing and the number of mumps cases and on translating this technique to other infectious diseases, such as measles, of which the number of cases is rising across Europe.

## Statements

### Ethical statement

This study was conducted in the context of outbreak control, as outlined in the Dutch Public Health Act. The study did not require approval from the medical ethical committee, as it does not concern medical scientific research as defined by the Dutch Central Committee on Research Involving Human Subjects. This was confirmed by the Medical Research Ethics Committee (METC NedMec).

### Use of artificial intelligence tools

None declared

### Data availability

Data is available from the corresponding author upon reasonable request.

### Authors’ contributions

MJ, AD, and LJ performed the outbreak investigation and drafted the manuscript, MJ and AD performed the data analysis, MG and GM were involved in laboratory analysis, and RS and GM performed local wastewater sampling and provided the passive samplers. MJ, AD, RS, GS, GM, and LJ designed the study protocol. All authors were involved in the interpretation of results, and revision of the manuscript.

## Acknowledgements

Genotyping was performed at the National Institute for Public Health and the Environment as part of its status as national reference laboratory. The authors extend special thanks to Putri Ayu Fajar for her work on the laboratory analyses.

